# Coronavirus-related online web search desire amidst the rising novel coronavirus incidence in Ethiopia: Google Trends-based infodemiology

**DOI:** 10.1101/2020.07.23.20158592

**Authors:** Behailu Terefe, Alessandro Rovetta, Asha K Rajan, Mengist Awoke

## Abstract

**Background:** During disease outbreaks, social communication and behaviors are very important to contain the outbreak. Under such circumstances, individual activities on online platforms will increase tremendously. This will result in the circulation useful or misleading/misinformation (infodemic monikers) in the community. Thus, exploring the online trending information is highly crucial in the process of containing disease outbreak. Therefore, this study aimed to explore users’ concerns towards coronavirus-related online web search activities and to investigate the extent of misleading terms adopted for identifying the virus in the early stage of COVID-19 spread in Ethiopia.

**Methods:** Google Trends was employed in exploring the tendency towards coronavirus-related web search activities in Ethiopia from March 13 to May 8, 2020. Keywords of the different names of COVID-19 and health-related issues were used to investigate the trends of public interest in searching from Google over time. Relative search volume (RSV) and Average peak comparison (APC) were used to compare the trends of online search interests. Pearson correlation coefficient was calculated to check for the presence of correlation.

**Result:** During the study period, “corona,” “virus,” “coronavirus,” “corona virus”, “China coronavirus,” and “COVID-19”, were the top names users adopted to identify the virus. In almost all search activities, the users’ employed infodemic monikers to identify the virus (99%). “Updates” related issues (APC=60, 95% CI, 55 – 66) were the most commonly trending health-related searches on Google followed by mortality (APC=27, 95% CI, 24 – 30) and symptoms (APC=55, 95% CI, 50 – 60) related issues. The regional comparison showed the highest cumulative peak for the Oromia region on querying health-related information from Google.

**Conclusion:** This study revealed an initial increase in the public interest of COVID-19 related Google search, but this interest was declined over time. Tremendous circulation of infodemic monikers for the identification of the virus was also noticed in the country. The authors recommend concerned stakeholders to work immensely to keep the public alert on coronavirus-related issues and to promote the official names of the virus to decrease the circulation of misleading and misinformation amid the outbreak.

## Background

Globally, the emerging of the COVID-19 pandemic is still at its rise affecting more people. The family of coronaviruses is already known for its spread of severe acute respiratory syndrome (SARS) in China and Middle East respiratory syndrome (MERS) in Saudi Arabia in 2003 and 2012, respectively [1]. Bats are believed to be the nidus for virus transmission, remaining as an intermediary zoonosis [2,3]. As compared to its peers, COVID-19 is a highly infectious disease, and worldwide nearly six million confirmed COVID-19 cases were recorded, as of late May 2020 [4].

Until May 22, 2020, COVID-19 infections were reported from 54 countries in Africa with cumulative confirmed cases of nearly a hundred thousand, and above three thousand deaths [5]. The first confirmed COVID-19 case in this continent was from Egypt and then Algeria [6,7]. In Ethiopia, the first COVID-19 case was detected and reported on March 13, 2020. Thereafter, incidences of COVID-19 cases in the country are increasing day to day and as of late May 2020 report, there were a total of 831 confirmed COVID-19 cases and 7 confirmed deaths [8].

During a disease outbreak, social communication and behaviors are believed to be just as important to public health as tests and diagnosis [9]. As a result, with the emerging spread of COVID-19, individual activity in almost all social media reaches its peak. Various studies on the mode of using social media for assessing knowledge, health promotion messaging and crisis communication has been carried out [10–12]. Additionally, the advancement in surveillance systems has enabled the scientific community to assess a large amount of real-time data globally which helps stakeholders to identify primary issues of concerns and address them. Sources for these real-time data include Google searches, Facebook, twitter posts, Wikipedia access, and Google plus [1,13].

The science which deals with the determinants and distribution of information in an electronic medium, in particular the internet sources, or in a population to inform public health and public policy is called infodemiology [14,15]. Google Trends is the most widely used website for infodemiologic studies of public health concern. It has advantages for analyzing, monitoring, forecasting, and nowcasting health topics related to infectious disease outbreaks and others [16,17]. There are about 526 million internet users in Africa and 4.2 million in Ethiopia [18]. According to the 2020 report, Africa’s internet penetration was estimated to be 39.3%, in Ethiopia, the estimated penetration was 17.8 % [19].

On online search metrics, like Google Trends, changes in the trends of information and communication on the internet can indicate changes in public health which can have a negative or positive impact. Especially during an outbreak, there will be a circulation of misinformation in the communities. Fake news, misleading, and misinformation circulating online in the community are referred to as infodemic monikers. Thus, checking the trends of information related to public health concerns is highly crucial, particularly during disease outbreak [13]. So far infodemiologic studies are limited in Africa, as well as in Ethiopia. As a result, this study explored users’ concern towards coronavirus-related online web searches and examined the extent of infodemic monikers adopted for identifying the virus in the country in the early stage of COVID-19 spread in Ethiopia.

## Methods

### Aim, design and setting

### Aim

The aim of this study was to explore users’ concerns towards coronavirus-related online web search activities and to investigate the extent of infodemic monikers adopted for identifying the virus in the early stage of COVID-19 spread in Ethiopia from March 13 to May 8, 2020.

In this study the term “infodemic monikers” were defined as the adoption of too generic names such as “coronavirus” or terms with ethnic associations like “china coronavirus” and “chinese coronavirus” or the use of separate terms “corona virus”, “virus”, “corona”(and the like). The use of this terms has caused very serious economic damage, have led to the use of information related to old coronaviruses not valid for the novel coronavirus, and episodes of ethnic-racism against people from China[20–23].

### Study design

Google Trends-based infodemiologic study

### Setting of the study

the study explored the trending information circulating in Ethiopia. Ethiopia has nine regions and two self-administrative cities [24].

### Description of search strategy

Google Trends was employed in exploring the online trending information. This website analyzes the types and trends of information searched from Google across different geographical areas and languages over a specific period. The website has features, such as explore, Trending Searches, and Year in Search, and it depicts graphs to compare the search volume interest over time for keywords entered. It also allows the user to compare the RSV of searches between two or more terms [18,25]. The search result from Google Trends ranges from 0 to 100, indicating very low to very high search volumes, respectively. The use of this website permits retrieval of queries for any keyword entered, and up to five groups of terms can be compared at a time. In our study, keywords for the different names of COVID-19 and health search-related issues associated with this virus circulating in Ethiopia were extracted from the headlines of the top known local news websites [26], regional media, and federal government health institutions’ (Ministry of health, and Ethiopian public health institute) websites [27,28]. Accordingly, the data were retrieved from Google Trends using these keywords in English and most spoken local languages in Ethiopia. Each of the extracted keywords was initially searched, and the resulting related-query were researched for further keywords to be included. After exhaustive identification of the different keywords used to refer to the COVID-19 and related health issues, keywords with the highest RSV was identified against which other terms were compared. During the analysis, health-related keywords with similar concepts were grouped together into the following classes: Updates, Mortality, Symptoms, Cures and Prevention (see Additional file 2). Updates-related searches were categorized as: world update (any coronavirus-related updates searches containing the term global, world, WHO) and local updates (any coronavirus-related update searches containing the country name Ethiopia or Unspecified update-related searches). RSV, APC, and graphs were used to compare the trends of online search interest over time for the keywords at a national level and across regions in the country.

## Data analysis

To determine the average trend of the various Google queries at the national and regional level, we used the APC index, which is the RSV mean value over the considered period. Each keyword APC has been presented with a 95% confidence interval. To do this, we calculated the standard error of the mean (SEM) as σ/√N, with σ standard deviation; then, we built the interval [APC-2×SEM, APC+2×SEM]. Furthermore, for all the regions whose data were accessible, we looked for any linear correlation between the number of COVID-19 cases per region and the various health-related searches. The Pearson coefficient expressing the above correlation was indicated with the letter “R” and reported with its p-value. We chose R=0 as H_0_null hypothesis (R ≠ 0 as H_1_ resp.) and α = .05 as a threshold value.

## Results

## Covid-19 identification search

The top names adopted by the Google users to identify the virus in Ethiopia were “corona,” “virus,” “coronavirus,” “corona virus”, “China coronavirus,” and “COVID-19” (Fig 1).

**Figure 1:**
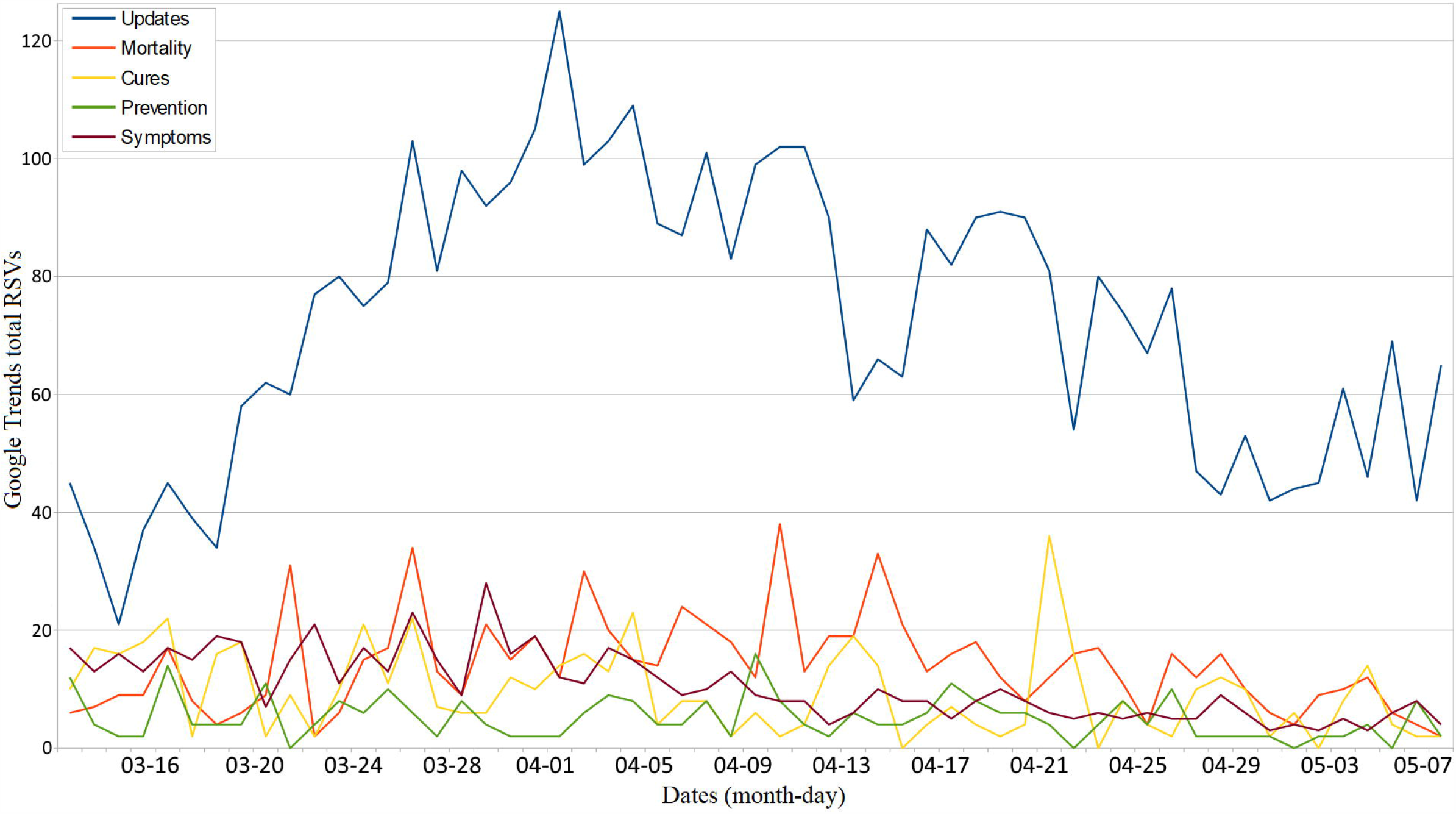
Shows the trends of the main names adopted by Google users to identify the virus in Ethiopia over the study period.

The overall trends starting from the first COVID-19 case report in Ethiopia showed the greatest relative search value for the keyword “corona” followed by “coronavirus,” with APC of 60 (95% CI, 55 – 66) and 55 (95% CI, 50 - 60), respectively. On April 06, the search for the keyword “corona” achieved the highest RSV (100), while the highest RSV (92) for the keyword “coronavirus” was on April 05, 2020. Starting from April 18 the searching trend was changed, and the term “coronavirus” became the predominantly searched term followed by the term “corona”. Of the keywords searched by the users to identify the virus, nearly all of them are infodemic monikers in type (overall relative search percentage of 99%) (Table 1).

**Table 1:** Shows in detail the APCs and the type of moniker used by users to identify the virus.

| **Keyword** | **APC** | **95% CI (SEM)** | **Peak RSV** | **Moniker type** | **Relative %** |
| --- | --- | --- | --- | --- | --- |
| **corona** | 60 | 55 – 66 | 100 | Infodemic | 35 |
| **Coronavirus** | 55 | 50 – 60 | 92 | Infodemic | 32 |
| **Virus** | 27 | 24 – 30 | 51 | Infodemic | 16 |
| **corona virus** | 26 | 23 – 29 | 47 | Infodemic | 15 |
| **China coronavirus\*** | 2 | 2 – 3 | 5 | Infodemic | 1 |
| **COVID-19** | 1 | 1 – 2 | 2 | Scientific | 1 |
| **other scientific terms** | 0 | 0 – 0 | 0 | Scientific | 0 |

\*group of words (see Additional file 1), APC: Average Peak Comparison, SEM: Standard Error of the Mean, RSV: Relative Search Volume

## Health-related search

The top five searches related to health in Ethiopia after the first COVID-19 case report were grouped under COVID-19 related “Updates,” “Mortality,” “Symptoms,” “Cures” and “Prevention”. On average, the keyword “updates”-related search was the highest of all searched term (a total APC of 72, 95% CI, 66 – 79) followed by “mortality” related (a total APC of 14, 95% CI, 12 – 16) and “symptoms” related (a total APC of 11, 95% CI, 9 – 12) queries (see Additional file 2). On April 02, 2020, the “updates” related searches in Ethiopia reached the highest (total RSV, 125). The “mortality” related searches reached a maximum peak (total RSV, 38) on April 10, 2020, whereas “Cures” related searches reached the highest peak (total RSV, 36) on April 22, 2020 (Table 2).

**Table 2:** Shows in detail the APCs and highest peak value of the health-related Google searches over the specified period.

| **Keyword group** | **Total APC** | **95% CI** | **Total RSV peak** | **Relative %** |
| --- | --- | --- | --- | --- |
| **Updates** | 72 | 66 – 79 | 125 | 65 |
| **Mortality** | 14 | 12 – 16 | 38 | 13 |
| **Symptoms** | 11 | 9 – 12 | 28 | 10 |
| **Cures** | 9 | 8 – 11 | 36 | 8 |
| **Prevention** | 5 | 4 – 6 | 16 | 5 |

APC: Average Peak Comparison, RSV: Relative Search Volume

The public desire towards health-related Google search over the specified period indicated an initial rise in the health-related search desire, but a decline in this interest was noticed over time. (Fig 2).

**Figure 2:**
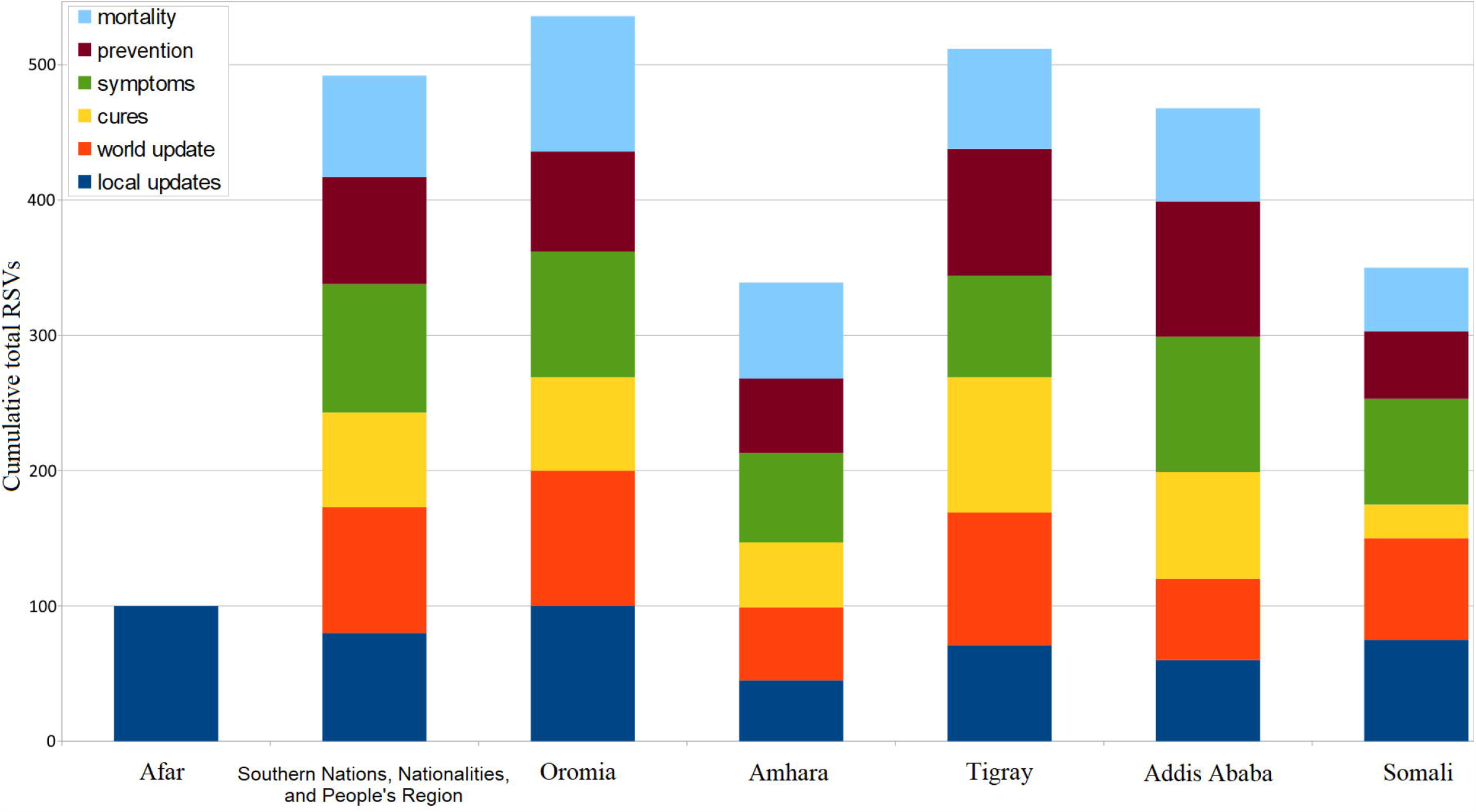
Shows trends of the health-related Google searches by the users over the specified period.

## Distribution of health-related cumulative Google searches across regions and cities

Distribution of health-related cumulative searches indicated the highest maximum peak search in the Oromia region (cumulative total RSVs, 536) followed by the Tigray (cumulative total RSVs, 512) and Southern Nations, Nationalities, and Peoples Region (cumulative total RSVs, 492) on searching health-related information (Fig 3).

**Figure 3:**
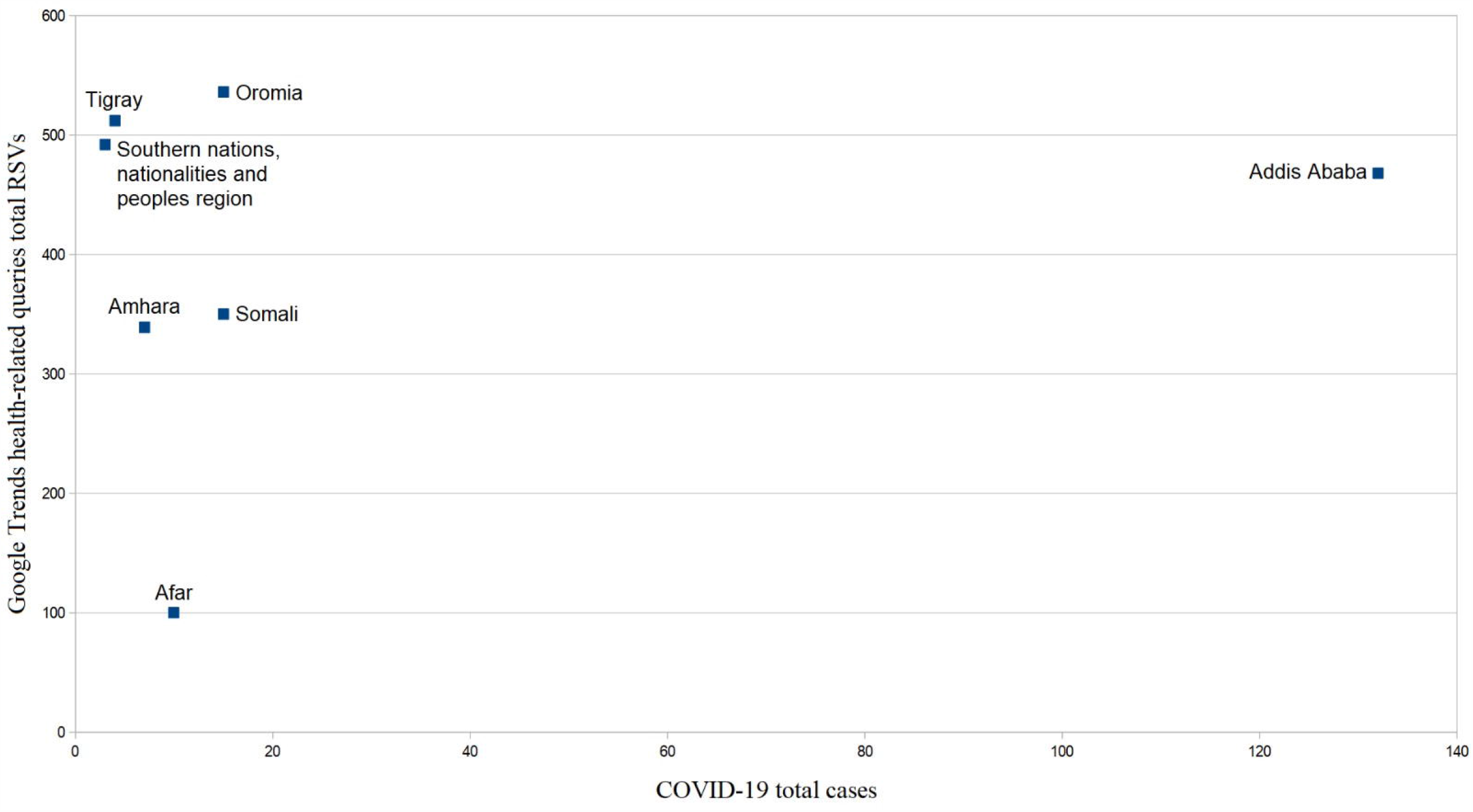
Shows the cumulative Google searches made to health-related searches over the specified period.

The frequently searched keyword was “local updates”; with the highest relative search percentage seen from Afar (93%), and the least from Addis Ababa (48%) (Table 3).

**Table 3:** Shows the relative percentage distribution of health searches by region/city over the specified period.

| **Region** | **local**<br>**updates %** | **world**<br>**update %** | **cures %** | **symptom %** | **precautions %** | **mortality %** |
| --- | --- | --- | --- | --- | --- | --- |
| **Afar** | 93 | 0 | 0 | 0 | 0 | 7 |
| **Oromia** | 63 | 5 | 7 | 5 | 9 | 11 |
| **SNNPR\*** | 67 | 5 | 9 | 6 | 5 | 7 |
| **Tigray** | 68 | 5 | 3 | 4 | 8 | 12 |
| **Somali** | 71 | 6 | 9 | 3 | 9 | 3 |
| **Addis**<br>**Ababa** | 48 | 9 | 14 | 9 | 9 | 10 |
| **Amhara** | 53 | 8 | 11 | 7 | 10 | 12 |

\*SNNPR: Southern Nations, Nationalities, and Peoples Region

The peak total RSV for the search term “local updates” related issue for Afar and Oromia region (total RSV, 100 each) was the highest, whereas for the“world update” related issue the highest total RSV was observed from Oromia (total RSV, 100) followed by Tigray region (total RSV, 98). The “cures” related issues RSV was highest for the Tigray region (total RSV, 100) and lowest for the Afar (total RSV, 0). Both the “symptoms,” and “prevention” related issue queries achieved the highest total RSV for Addis Ababa (total RSV, 100 each), while “mortality” related issue search was the highest in the Oromia region (total RSV,100) followed by Southern Nations, Nationalities, and Peoples Region (total RSV, 75) and Tigray region (total RSV,74). In the correlation test between the Google health searches and COVID-19 cases per region, all p-values confirmed the null hypothesis i.e. no significant linear correlation was detected (Table 4).

**Table 4:**
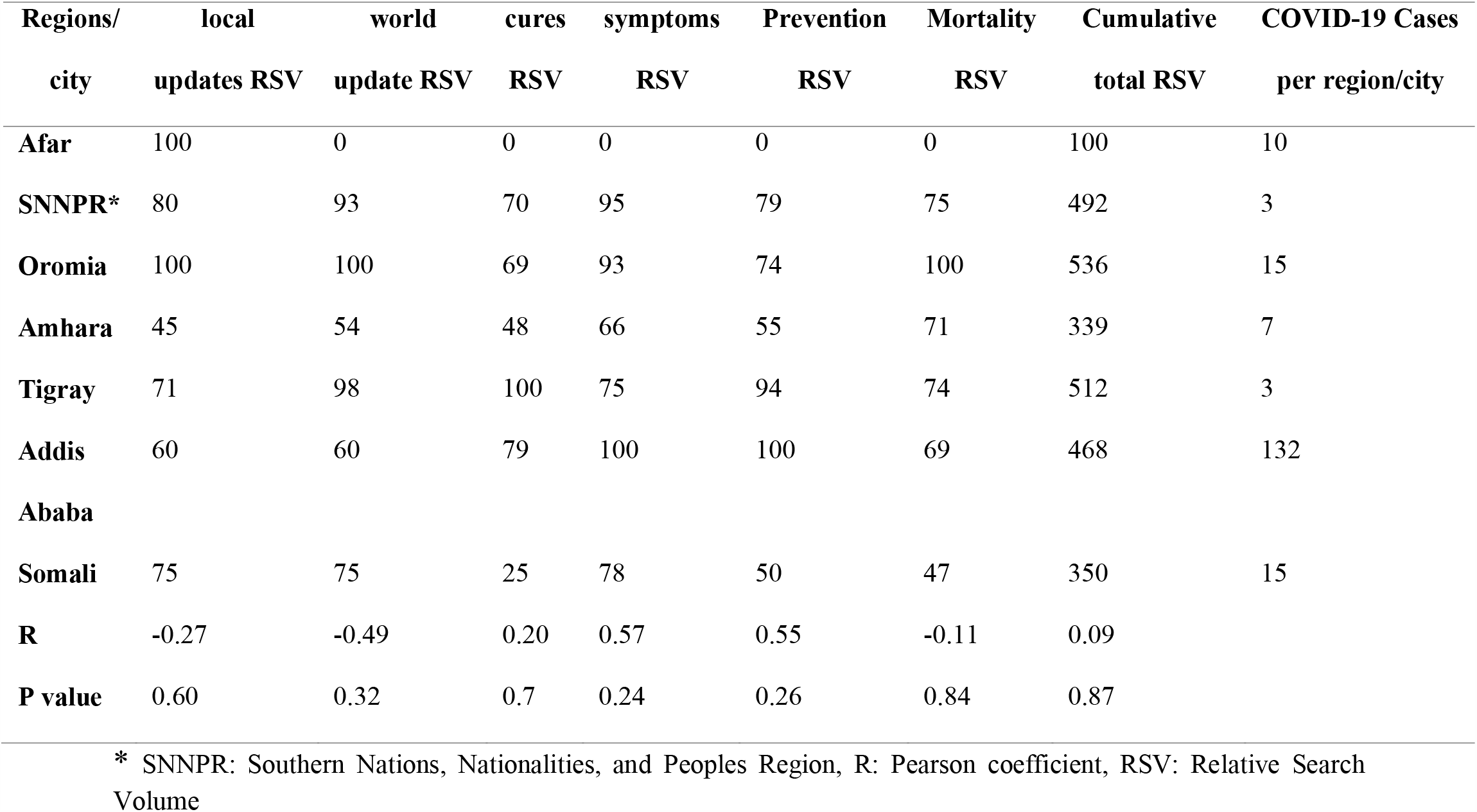
Shows the volume of health queries of each region/city over the specified period with the correlation test results.

| Regions/<br>city | local<br>updates RSV | world<br>update RSV | cures<br>RSV | symptoms<br>RSV | Prevention<br>RSV | Mortality<br>RSV | Cumulative<br>total RSV | COVID-19 Cases<br>per region/city |
| --- | --- | --- | --- | --- | --- | --- | --- | --- |
| **Afar** | 100 | 0 | 0 | 0 | 0 | 0 | 100 | 10 |
| **SNNPR\*** | 80 | 93 | 70 | 95 | 79 | 75 | 492 | 3 |
| **Oromia** | 100 | 100 | 69 | 93 | 74 | 100 | 536 | 15 |
| **Amhara** | 45 | 54 | 48 | 66 | 55 | 71 | 339 | 7 |
| **Tigray** | 71 | 98 | 100 | 75 | 94 | 74 | 512 | 3 |
| **Addis**<br>**Ababa** | 60 | 60 | 79 | 100 | 100 | 69 | 468 | 132 |
| **Somali** | 75 | 75 | 25 | 78 | 50 | 47 | 350 | 15 |
| **R** | -0.27 | -0.49 | 0.20 | 0.57 | 0.55 | -0.11 | 0.09 |  |
| **P value** | 0.60 | 0.32 | 0.7 | 0.24 | 0.26 | 0.84 | 0.87 |  |

\* SNNPR: Southern Nations, Nationalities, and Peoples Region, R: Pearson coefficient, RSV: Relative Search Volume

The distribution of COVID-19 cases across the regions/city versus the total Google Tends health-related searches is depicted in the scatter plot below (Fig 4).

**Figure 4:**
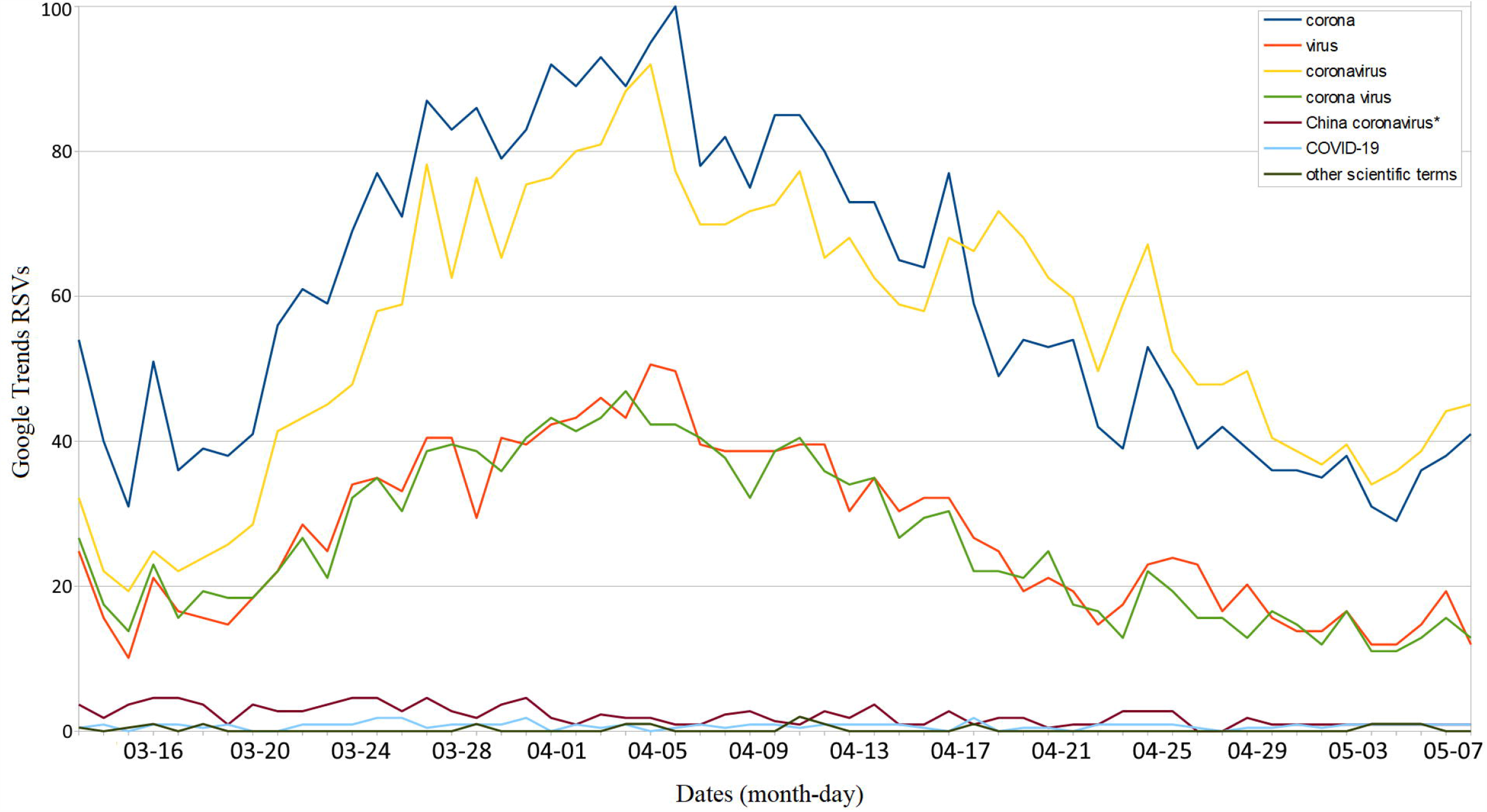
Scatter plot showing the distribution of COVID-19 cases across the regions/city versus the total Google Tends of health-related searches interest over the specified period.

## Discussion

In this explorative infodemiology study, we tried to investigate the trend of online search behaviors towards coronavirus in the early stage of COVID-19 spread in Ethiopia using Google Trends. We have also attempted to determine the extent of infodemic monikers used for identifying the virus in the country. On average the term “corona” was the predominantly adopted name in the community for identifying the virus, and the keyword “updates” related search was the highest of all health searches on Google throughout the study period. In almost all searches to identify the virus from Google, the users adopted infodemic monikers. Distribution of health-related cumulative searches indicated the highest maximum peak search in the Oromia region as compared to the others. No significant correlation was noted between the Google health searches and the number of COVID-19 cases per region.

The social contagion of infodemic monikers (erroneous, misleading, and misinformation) during the COVID-19 outbreak has affected global efforts to fight the epidemic. This has resulted in the high demand for timely and trustworthy information about COVID-19 during the outbreak [29]. In our study, 99% of the keywords used in Google search to identify the virus were infodemic. These terms are either they will not specifically identify the COVID-19 (these are keywords: “corona”, “Coronavirus”, “Virus” and “corona virus”) or they are stigmatic ascribing the virus to a specific ethnic group (China coronavirus). They are prone to transmit inappropriate information in the community. Provided that most of the horns of African countries, including Ethiopia are predicted to have a severely alarming epidemic prognosis, the circulation of these misleading and misinformation will probably impose a significant impact in the process of containing COVID-19 in this region [30].

Looking at the trends of the virus name adopted for search in Google, on April 06, 2020, the keyword “corona” has achieved the highest peak search. This happened one day after the first COVID-19 related death report in Ethiopia [31,32]. On this day, the global reports of confirmed COVID-19 cases were above 1 million [4]. On the other hand, April 02, 2020, marked the highest “updates” related Google searches in Ethiopia. This was the day that marked above one million confirmed COVID-19 cases, worldwide [8]. Navigation of health-related cumulative Google search distribution across regions/city in Ethiopia indicated the highest maximum peak search in the Oromia region followed by the Tigray region on searching health-related information. In Ethiopia, the first COVID-19 case was reported in the Adama city, Oromia region`s capital. This has led to the early complete ban of public transportation systems in the region [33]. The Tigray region was also the first to declare a 15-day region-wide state of emergency, like banning all travel and public activities including, closure of all cafes, restaurants, and banning of landlords from evicting tenants or increasing rent. Additionally, any travelers entering the region were also required to report to the nearest health offices [34,35]. These scenarios might have alerted the public in both regions to highly desire online information towards COVID-19 related health issues.

## Conclusions and recommendations

This study indicated an initial increase in the interest of COVID-19 related Google search, but this interest was declined after some time. Tremendous circulation of infodemic monikers related to identifying the virus was also noticed in the country. Furthermore, numbers of COVID-19 cases across the regions/city was not found to have a significant association with the highest peak health-related searches. Based on the findings, the authors of this study recommend the government and other stakeholders to work immensely to keep the public alert on coronavirus-related issues. Additionally, the official names of the virus, like “COVID-19”, “SARS-CoV-2” have to be promoted in the public to decrease the circulation of misleading and misinformation amid the outbreak.

## Data Availability

All data are available and included in the manuscript and supplementary materials.

## List of abbreviation and acronyms

APC: Average Peak Comparison
CI: Confidence Interval
MERS: Middle East Respiratory Syndrome
R: Pearson correlation coefficient
RSV: Relative Search Volume
SARS-COV-2: Severe Acute Respiratory Syndrome Coronavirus-2
SEM: Standard Error of the Mean
SNNPR: Southern Nations, Nationalities, and Peoples Region

## Declarations

### Ethics approval and consent to participate

Not applicable

### Consent for publication

Not applicable

### Availability of data and materials

The dataset supporting the conclusions of this article is included within the article and its additional file (see Additional file 3).

### Authors’ contributions

BT, AR, AKR and MA: Conceptualization, Data curation, Formal analysis, Investigation, Supervision, Validation, Visualization, Writing – original, draft preparation & editing. All authors have read and approved the manuscript.

## Acknowledgements

We declare no acknowledgment for any third party as all the activities were carried-out by the authors.

## Supplementary files

**Additional file 1**: keywords having the same concept and categorized together to identify the virus name related queries

**Additional file 2:** keywords having nearly the same concept and categorized together to identify the health-related users’ queries

**Additional file 3:** Dataset .csv file: Coronavirus-related online web search desire amidst the rising novel coronavirus incidence in Ethiopia: google trends-based infodemiology

